# Perceptions of aging well among older adults with heart failure: insights from a qualitative study

**DOI:** 10.64898/2026.06.11.26355065

**Authors:** Ro-Jay Reid, Deborah Ofosu, Parag Goyal, Atami De Main, W. Marcus Lambert, Iris Navarro-Millán, Madeline R. Sterling, Mangala Rajan, Orysya Soroka, Monika M. Safford

## Abstract

**Background:** Heart failure (HF) is a prevalent and often debilitating cardiovascular condition among older adults, frequently accompanied by multimorbidity, functional limitations, and the need to age in place. Traditional models of successful aging emphasize disease absence and preserved function, yet most individuals with HF live with ongoing symptoms and chronic health challenges. How older adults with HF define aging well, particularly across different socioeconomic contexts, remains underexplored.

**Objectives:** To explore how older adults with HF conceptualize aging well and to identify perceived facilitators and barriers across more and less resourced New York City neighborhoods.

**Methods:** We conducted semi-structured interviews with 20 adults diagnosed with HF residing in Manhattan and Brooklyn neighborhoods classified by 2019 United States Census data. Interviews were guided by Rowe and Kahn’s model. Transcripts were analyzed using an inductive-deductive thematic approach and interpreted in alignment with the Healthy People 2030 framework.

**Results:** Participants had a mean age of 69 years; 50% identified as Black and 50% were women. Despite functional limitations, 65% reported aging well. Five themes emerged: maintaining physical function, maintaining cognitive function, sustaining social relationships, avoiding pain, and promoting overall well-being. Avoiding pain and promoting well-being extended beyond traditional models. Neighborhood context shaped priorities, with financial stability emphasized in more affluent areas and social cohesion prioritized in less affluent communities.

**Conclusions:** Older adults with HF frequently perceive themselves as aging well despite chronic illness, reframing successful aging beyond disease avoidance. These findings support a patient-centered, place-informed model of aging well with implications for healthcare delivery and policy.

## INTRODUCTION

Heart failure (HF) is a prevalent and often debilitating cardiovascular condition among older adults, frequently accompanied by multimorbidity, functional limitations, and the need to age in place [1,2]. As most older adults live with chronic illness, there is increasing interest in understanding how aging well is experienced despite ongoing health challenges. HF provides a particularly important context for this inquiry, given its substantial symptom burden and impact on daily functioning.

Rowe and Kahn’s Successful Aging framework has strongly shaped gerontological research, defining successful aging as low probability of disease/disability, high cognitive and physical functioning, and active engagement in life [2,3]. While influential, the model gives less attention to subjective experiences and broader social and environmental contexts [5–7]. In response, scholars have expanded the concept toward broader frameworks such as aging well, which retain elements of function and engagement but emphasize psychosocial adaptation, resilience, purpose, and overall well-being [8]. These perspectives acknowledge that aging well may coexist with chronic illness. However, limited qualitative research has examined how older adults with HF define aging well especially across different socioeconomic and neighborhood contexts [9–11].

Aging well is shaped not only by individual health status but also by the environments in which people live [6,7]. Neighborhood context influences access to health-promoting resources, opportunities for social engagement, and conditions that support or constrain physical activity. Differences in neighborhood socioeconomic status may amplify or buffer health risks through variation in safety, walkability, healthcare access, and social infrastructure, contributing to disparities in physical, cognitive, and emotional well-being [12,13]. These contextual influences align with the World Health Organization (WHO) call for age-friendly environments [14].

To address these gaps, we conducted a qualitative study of older adults living with HF in affluent and less affluent New York City neighborhoods. We focused on HF because it is a high-mortality cardiovascular condition characterized by progressive functional decline, substantial symptom burden, and shortened life expectancy, requiring patients to confront serious illness while continuing to age in place [1,2,10]. In addition, HF is frequently associated with reduced mobility, making neighborhood context especially relevant to daily functioning, access to care, and social engagement [1,2,10]. For these reasons, HF provides a particularly informative lens through which to examine how aging well is defined in the setting of chronic cardiovascular disease. Our objective was to examine how individuals with HF define aging well and to identify perceived barriers and facilitators shaped by neighborhood context. By centering patient perspectives, this study seeks to inform more patient-aligned and context-sensitive models of HF care.

## METHODS

### Study Design and Participants

This phenomenological, cross-sectional qualitative study used semi-structured interviews with a purposive sample of 20 adults with a physician diagnosis of HF. Participants were recruited from New York-Presbyterian Hospital Weill Cornell Internal Medicine Associates and the Heart Failure with Preserved Ejection Fraction Program at Weill Cornell Medicine.

Inclusion criteria were HF diagnosis for at least one-year, English fluency, and willingness to participate in a telephone or in-person interview. Exclusion criteria included physician-diagnosed dementia, hospice residence, active cancer treatment, or residence in a nursing home.

Participants were recruited from both lower-income and more affluent neighborhoods in Manhattan and Brooklyn. Neighborhood status was determined using 2019 American Community Survey zip code-level data. Zip codes were categorized into tertiles based on the percentage of residents living below the federal poverty threshold: 0-11.7%, 11.8-20.7%, and 20.8-36.8%. Participants residing in zip codes with 0-11.7% of residents below the poverty line were classified as living in more affluent neighborhoods, whereas those in zip codes with 11.8-36.8% were classified as living in less affluent neighborhoods.

The study was approved by the Biomedical Research Alliance of New York and the Weill Cornell Medicine Institutional Review Board. Verbal informed consent was obtained from all participants in accordance with federal human subjects’ regulations. Participants received a $50 gift card upon completion.

### Data Collection

The study was guided by the Consolidated Criteria for Reporting Qualitative Research [15]. One investigator (RR) conducted semi-structured telephone interviews between March 6 and October 31, 2023.

A pilot-tested interview guide, modeled after Rowe and Kahn’s theoretical framework [3,4], was used to facilitate discussion (see Supporting material). Example interview questions are presented in Table 1.

**Table 1.**
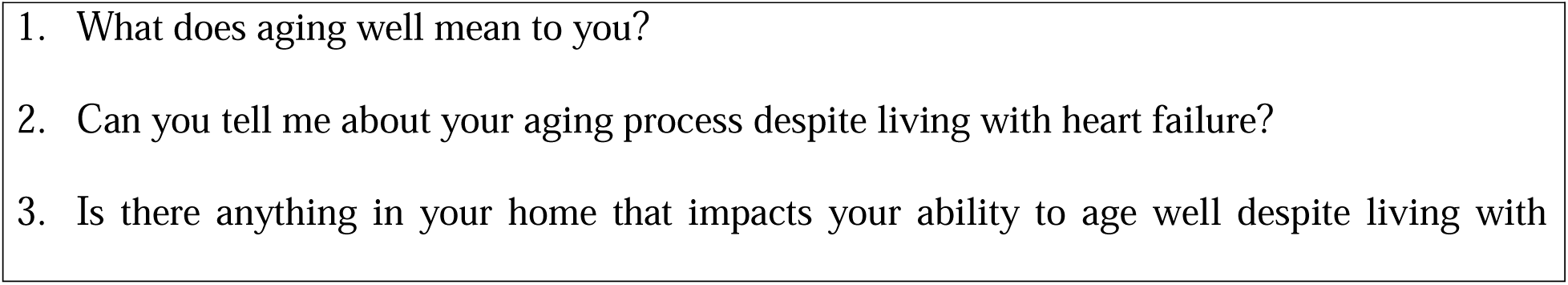
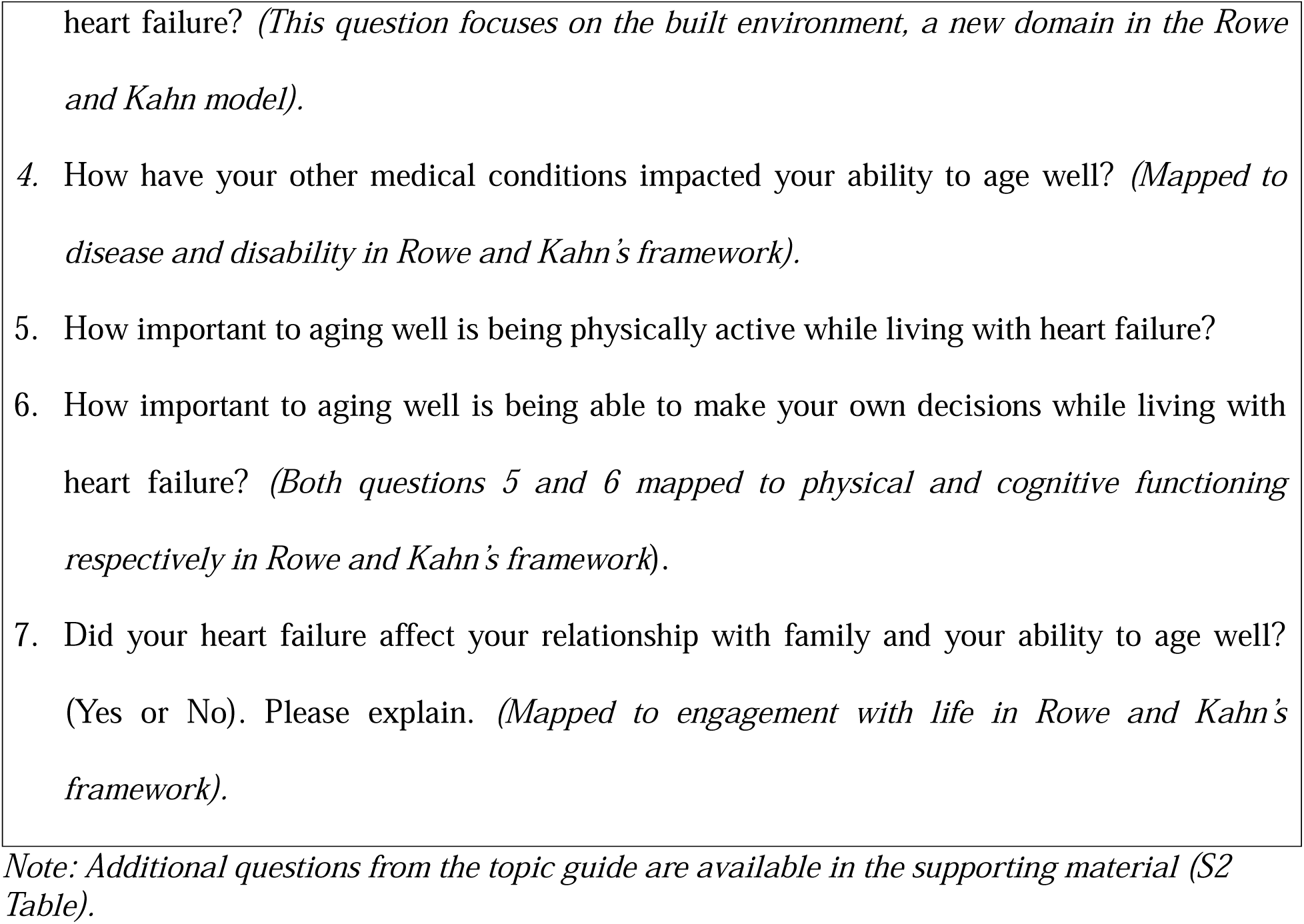
A sample of topic guide questions and their alignment with Rowe and Kahn’s framework for successful aging.

At the conclusion of each interview, participants completed a brief demographic survey. Medical records were reviewed to confirm functional status using the New York Heart Association (NYHA) classification, number of medications, year of HF diagnosis, and comorbid conditions. Interviews lasted approximately 30-60 minutes, were audio-recorded, and transcribed verbatim.

### Data Analysis

Audio recordings were transcribed verbatim and verified for accuracy by Production Transcription Inc., in accordance with Health Insurance Portability and Accountability Act regulations and institutional guidelines. Data were managed and analyzed using MAXQDA version 24.7, a qualitative data analysis software.

The first and second authors (RR and DO) independently coded three transcripts using thematic analysis, a method appropriate for health-related qualitative research guided by a conceptual framework [16,17]. Coding was informed by Rowe and Kahn’s framework [3,4] while remaining open to inductively derived concepts. Following independent coding, discrepancies were reconciled through discussion, resulting in development of an initial codebook [18,19]. The refined codebook was applied to the remaining transcripts, with new codes incorporated through an iterative deductive and inductive process [20]. Following the “stopping criterion” outlined by Francis et al. [21], saturation was reached after analysis of 15 interviews, when no new codes or substantive themes were identified. Five additional interviews were conducted and analyzed to confirm saturation; no new themes emerged, supporting the adequacy of the final sample size.

When disagreements arose during coding, excerpts were revisited until consensus was achieved. Codes or candidate themes that could not be reconciled were excluded from the final thematic structure [22], consistent with established qualitative analytic approaches and reflecting a deliberate focus on themes with sustained interpretive support across coders and transcripts. Coders maintained analytic memos and engaged in reflexive discussions regarding how their clinical and research perspectives might influence interpretation, ensuring that themes remained grounded in participants’ narratives [17].

Codes were grouped into categories and refined into overarching themes. To facilitate comparison of contextual influences across neighborhood groups, we developed a heatmap summarizing the relative prominence of facilitators and barriers to aging well according to the Healthy People 2030 Social Determinants of Health framework [23]. This framework was selected because it provides a nationally recognized structure for organizing social, economic, and environmental factors that influence health and well-being and aligns with the study’s focus on neighborhood-level influences on aging well. Following theme development, coded excerpts were reviewed and classified as representing either a facilitator or barrier. For each domain, frequencies were calculated as the proportion of participants within each neighborhood group who endorsed the theme at least once during their interview. Thus, heatmap values reflect participant-level endorsement rather than the number of times a theme was mentioned. Color intensity was used to visually represent the relative prevalence of themes across neighborhood groups, with darker shading indicating a greater proportion of participants endorsing a given domain.

Intercoder reliability was assessed using Cohen’s kappa to evaluate agreement beyond chance and enhance analytic rigor [24]. Additional analytic details are provided in the Supplemental Material.

Representative quotations were selected to illustrate major themes while balancing commonly expressed perspectives with illustrative insights.

## RESULTS

### Sample characteristics

A total of 20 interviews were conducted. Saturation was achieved after 15 interviews, with five additional interviews completed to confirm code saturation and ensure equal representation of participants from more affluent and less affluent neighborhoods (10 participants in each group). The final coding structure demonstrated a high level of agreement between coders, with a Kappa statistic of 84.4%. Participants ranged age from 40’s to 80’s years (mean 69). Half identified as women and half as White. Black participants were disproportionately represented in less affluent neighborhoods. Compared with participants in affluent neighborhoods, those in less affluent neighborhoods had lower educational attainment, lower income, poorer self-rated health, and greater functional impairment as it relates to the NYHA classification (Table 2). Despite these differences, most participants in both groups reported that they had aged well despite living with HF.

**Table 2:** Characteristics of participants by neighborhood affluence.

|  | **Overall** | **More affluent**<br>**Neighborhoods** | **Less affluent**<br>**Neighborhoods** |
| --- | --- | --- | --- |
|  | **n=20** | **n=10** | **n=10** |
| **Female %** | 50 | 50 | 50 |
| **Mean age in years (SD)** | 69.2 (12.0) | 72.9 (10.8) | 65.4 (12.4) |
| **Black or African American %** | 50 | 20 | 80 |
| **Marital Status %** |  |  |  |
| Married | 30 | 30 | 30 |
| Divorced | 10 | 10 | 10 |
| Single/Widow | 60 | 60 | 60 |
| **Educational attainment** |  |  |  |
| High school graduate or less | 40 | 20 | 60 |
| **Monthly Income %** |  |  |  |
| <=\$4000 | 60 | 50 | 70 |
| >\$4000 | 20 | 30 | 10 |
| Declined to report | 20 | 20 | 20 |
| **Employment Status %** |  |  |  |
| Employed | 15 | 20 | 10 |
| Unemployed | 30 | 10 | 50 |
| Retired | 55 | 70 | 40 |
| **Self-Rated Health %** |  |  |  |
| Excellent | 5 | 10 | 0 |
| Very good | 25 | 40 | 10 |
| Good | 25 | 20 | 30 |
| Fair | 30 | 30 | 30 |
| Poor | 15 | 0 | 30 |
| **Health Behaviors %** |  |  |  |
| Current Cigarette Smoking | 20 | 20 | 20 |
| Adherent to HF Medications | 95 | 100 | 90 |
| Adherent to low salt diet | 50 | 50 | 50 |
| Adherent to fluid restriction | 10 | 20 | 0 |
| **NYHA Classification %** |  |  |  |
| I | 40 | 50 | 30 |
| II | 30 | 30 | 30 |
| III | 30 | 20 | 40 |
| IV | 0 | 0 | 0 |
| **Number of Medications %** |  |  |  |
| 0 - 5 | 25 | 40 | 10 |
| 6 - 10 | 40 | 30 | 50 |
| ≥ 11 | 35 | 30 | 40 |
| **Years since HF diagnosis %** |  |  |  |
| 1 - 2 | 45 | 40 | 50 |
| 3-5 | 30 | 20 | 40 |
| ≥6 | 25 | 40 | 10 |

| **Medical Conditions %** |  |  |  |
| --- | --- | --- | --- |
| Hypertension | 85 | 100 | 70 |
| Type 2 diabetes | 30 | 20 | 40 |
| COPD or asthma | 20 | 10 | 30 |
| Cancer | 30 | 20 | 40 |
| Osteoarthritis | 35 | 50 | 20 |
| Chronic kidney disease | 40 | 20 | 60 |
| Coronary artery disease | 35 | 40 | 30 |
| Stroke or TIA | 10 | 0 | 20 |
| Atrial fibrillation | 35 | 20 | 50 |
| Depression | 25 | 40 | 10 |
| **Considers that they have aged well %** | 65 | 70 | 60 |

*Note: COPD = chronic obstructive pulmonary disease; HF = heart failure; NYHA = New York Heart Association; SD = standard deviation; TIA = transient ischemic attack.*

### Conceptualization of aging well despite HF

We identified five overarching themes that defined aging well for these participants, including: (1) maintaining physical functioning (2) maintaining cognitive functioning, (3) engaging in social connections, (4) avoiding pain and (5) achieving well-being, as illustrated in Fig 1.

**Fig 1:** Person-centered model of aging well despite living with heart failure. High Physical Function - Ability to carry out daily and physical activities independently. High Cognitive Function - Preservation of memory, attention, and decision-making abilities. Avoidance of Pain - Reduced or managed chronic pain to support mobility and comfort. Wellbeing - A sense of purpose, emotional balance, and mental health. Social Engagement - Participation in meaningful relationships and community activities.

#### Maintaining physical function

Participants described aging well as maintaining mobility, even when durable medical equipment (e.g., cane, walker, shower chair) was required. Physical function was framed not as freedom from disability, but as the ability to remain engaged in meaningful daily activities through adaptation and assistive strategies.

> “I use a walker now. I have what’s called a shower chair. So, I’m not like I used to be in terms of activities, but again, I don’t feel like that in specific makes me ill.”
>
> Woman, in her 50’s. Less affluent neighborhood.

#### Maintaining cognitive function

Maintaining cognitive function was described as essential for decision-making. Participants emphasized mental clarity as critical for managing responsibilities and preserving autonomy.

> “For me, aging well means staying mentally sharp…managing all my responsibilities…driving…keep up my computer skills.”
>
> Woman, in her 70’s. More affluent neighborhood.

#### Engagement in social connections

Social connections were central to aging well, particularly as sources of emotional support. Participants emphasized the importance of relationships in sustaining quality of life across neighborhood contexts.

> “Aging well means to be in a good relationship.”
>
> Man, in his 70’s. Less affluent neighborhood.

#### Avoiding pain

Avoidance of pain emerged as a crucial aspect of aging well while living with HF. Participants noted the challenges associated with managing pain, particularly given their heart condition.

> “I think aging well for me would include the absence of chronic pain, having sufficient capacity, physical, and mental capacity, to be reasonably mobile and enjoy the pleasures of social activity, whether it’s going to museums or theater or visiting family and friends or having them visit me.” Man, in his 70’s. More affluent neighborhood.
>
> “Well, I have arthritis all over my body. I’ve had two hip replacements, and I have been strongly advised to have knee replacements, which I choose not to do. I have no feeling in either of my feet, and to some degree, painful neuropathy in both feet. So, I find the level of pain I deal with is pretty high, and I do what I can to alleviate that. I take Advil. Because of my heart situation, many of the pain medications like meloxicam other things are not really advisable for me. But I do enjoy drinking, and I have a couple of glasses of wine at night, which help me to relax and not feel pain.”
>
> Woman, in her 70’s. More affluent neighborhood.

#### Achieving well-being

Well-being reflected participants’ ability to continue living in alignment with their preferred lifestyle, roles, and sense of self despite HF.

> “I believe in quality of life. So, aging well means that I can continue living the way I live, doing what I do, being able to handle all my responsibilities to myself and my home, and still looking chickie-chickie.”
>
> Woman, in her 80’s. More affluent neighborhood.

### Reasons for not aging well with HF

Seven of the twenty participants reported that they had not aged well. Across neighborhoods, reasons included early onset of chronic illness and functional decline. Participants in more affluent neighborhoods emphasized inability to exercise and cumulative health behaviors, whereas those in less affluent neighborhoods additionally described depression, obesity, and social and economic stressors.

> “No I have not aged well because I shouldn’t be having a lot of these things. My thing started at 34. I shouldn’t be having all these things that a senior citizen should be having. It’s ridiculous. But I was doing my life like I wanted to live my life, so (sighs) unfortunately, I am sort of paying the price. So, that’s not aging gracefully.”
>
> Woman, in her 50’s. More affluent neighborhood.
>
> “I have not aged well because my heart failure has affected everything - social life, working, which I would like to go back to work! Everything is not working out.”
>
> Woman, in her 60’s. Less affluent neighborhood.

### Facilitators and barriers to aging well

The facilitators and barriers to aging well aligned with the domains of the Healthy People 2030 framework [23] - education access and quality, economic stability, healthcare access and quality, neighborhood and built environment, and social and community context is highlighted in Table 3.

**Table 3.**
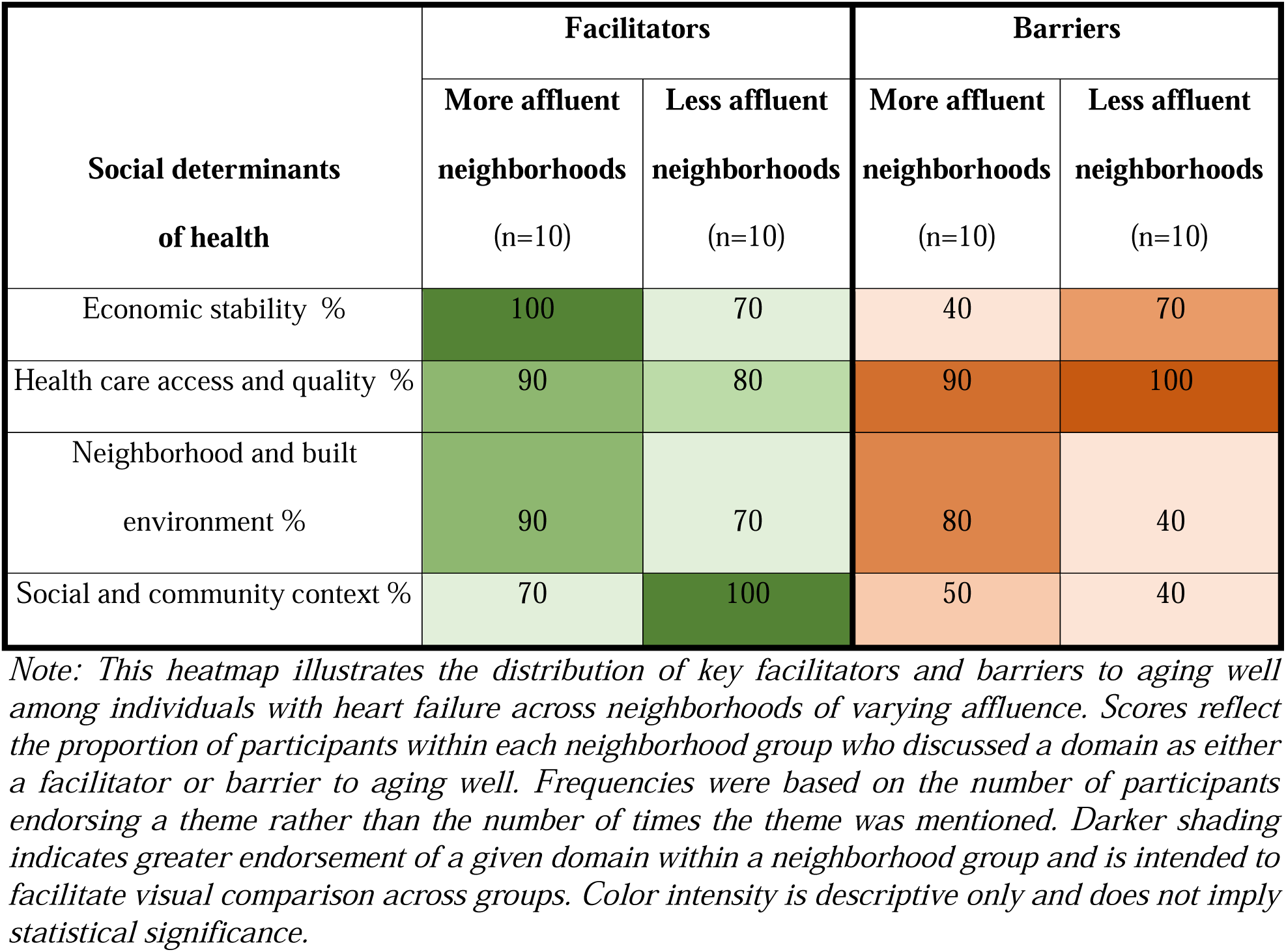
Heatmap of participant-endorsed facilitators and barriers to aging well with heart failure across neighborhood affluence groups.

| Social determinants<br>of health | Facilitators |  | Barriers |  |
| --- | --- | --- | --- | --- |
|  | More affluent<br>neighborhoods<br>(n=10) | Less affluent<br>neighborhoods<br>(n=10) | More affluent<br>neighborhoods<br>(n=10) | Less affluent<br>neighborhoods<br>(n=10) |
| Economic stability % | 100 | 70 | 40 | 70 |
| Health care access and quality % | 90 | 80 | 90 | 100 |
| Neighborhood and built<br>environment % | 90 | 70 | 80 | 40 |
| Social and community context % | 70 | 100 | 50 | 40 |

*Note: This heatmap illustrates the distribution of key facilitators and barriers to aging well among individuals with heart failure across neighborhoods of varying affluence. Scores reflect the proportion of participants within each neighborhood group who discussed a domain as either a facilitator or barrier to aging well. Frequencies were based on the number of participants endorsing a theme rather than the number of times the theme was mentioned. Darker shading indicates greater endorsement of a given domain within a neighborhood group and is intended to facilitate visual comparison across groups. Color intensity is descriptive only and does not imply statistical significance.*

#### Facilitators

In affluent neighborhoods, economic stability was the most frequently cited facilitator. In less affluent neighborhoods, social and community support emerged as the primary facilitator, often compensating for financial and healthcare challenges.

> “I mean, you probably understand from what I’ve said before that I’m pretty comfortable financially. If I thought of something that would be supportive, if I thought it would be useful to have it close at hand, I’ve pretty much been able to get it.”
>
> Man, in his 70’s. More affluent neighborhood.
>
> “If I don’t have the money to buy, and if they [nephews] have it, they get it for me. That’s the way it is. So, I don’t really have to be stressed out. I let them know or either they’ll call and see if I have this and have that. What I like about it is that my family, my friends and my neighbors are all like that. I’m surrounded with people who cares.”
>
> Woman, in her 80’s. Less affluent neighborhood.

#### Barriers

Across neighborhoods, healthcare access and quality was the most consistently cited barrier.

> “We all have problems just going out because of the Access-A-Ride. That system changed on us, and so it’s hard to get it sometimes. Even our doctors beginning to notice that work with us, because sometime we don’t get there on time, and they are limited to take so many patients a day, and sometime we have to make appointments like two or three times before they get us there, and sometime they won’t show up, and then when they do show up they could show up at the time we’re supposed to be at our appointments. That’s one reason it stress all of us out. It really does.”
>
> Woman, in her 80’s. Less affluent neighborhood.
>
> “I mean, after my last hospitalization I’ve been pretty determined and disciplined about trying to restore my capacity for regular physical exercise. It didn’t happen right away. It took a while to recover from the deterioration of muscles from the hospitalization experience.”
>
> Man, in his 70’s. More affluent neighborhood.

## DISCUSSION

Despite living with a chronic, life-limiting cardiovascular condition, most adults with HF perceived themselves as aging well. Aging well was conceptualized as a multidimensional process encompassing five interrelated domains: maintaining physical function, preserving cognitive function, sustaining social connections, avoiding pain, and achieving overall well-being. These findings are consistent with the WHO healthy aging framework [25] and extend traditional successful aging models [3–8,26–28] by demonstrating that aging well can coexist with symptomatic cardiovascular disease. While participants reaffirmed the importance of physical, cognitive, and social functioning, they also emphasized pain control and overall well-being - two domains often underrecognized in clinical practice [29,30] - as central determinants of whether they perceived themselves as aging well.

Pain emerged as a critical yet underrecognized barrier to aging well in HF. Our findings suggest that inadequately managed pain may contribute to maladaptive coping behaviors with potential cardiovascular consequences. In this study, one participant reported using alcohol while receiving anticoagulation for atrial fibrillation and occasionally taking nonsteroidal anti-inflammatory drugs for osteoarthritis to manage chronic pain. This example underscores the intensity of unmet symptom burden and highlights how self-directed coping strategies may increase risk for arrhythmias, bleeding complications, and HF decompensation - well-recognized complications in this population [31]. Pain management in older adults with cardiovascular disease is often complicated by concerns about polypharmacy, medication related adverse effects, and misconceptions that pain is an inevitable part of aging [32–34]. However, inadequate pain control may not only undermine independence and social engagement but may also worsen HF through physiologic mechanisms such as sympathetic nervous system activation and systemic inflammation [35]. These observations highlight the importance of healthcare providers assessing not only ischemic or chest pain but also uncontrolled musculoskeletal and chronic pain that may contribute to hospitalization risk or HF exacerbation. Collectively, these findings support proactive, multidisciplinary approaches to pain management, including nonpharmacologic strategies tailored to individuals living with HF [36–38].

Another novel dimension of this patient-centered model of aging well is well-being. Psychologists often conceptualized well-being as encompassing both hedonic well-being, which emphasizes happiness and life satisfaction, and eudaimonic well-being, which emphasizes purpose in life, autonomy, self-acceptance, personal growth, and the ability to live in accordance with one’s values. [5,39–42]. Consistent with these frameworks, participants described aging well in terms of maintaining independence, control over bodily functions, and overall quality of life despite living with heart failure. These factors closely align with dimensions of Ryff’s model of psychological well-being, particularly autonomy, environmental mastery, and self-acceptance [43]. Participants who perceived themselves as aging well often described adapting to age and disease-related limitations in ways that preserved independence and identity, reflecting principles of compensation and adaptation described in Selective Optimization with Compensation theory [28,29]. Together, these findings suggest that well-being reflects more than the absence of symptoms; it encompasses an individual’s ability to maintain a sense of self and purpose while adapting to changing capacities. However, because our interview guide did not specifically probe constructs such as purpose in life, personal growth, or self-actualization, future studies are needed to better understand how these dimensions contribute to perceptions of aging well. Multidimensional measures may therefore be needed to adequately capture this important domain of patient-centered aging well.

Lastly, our findings highlight the critical role of neighborhood, built environment and social context in aging well. As physical limitations progressed, features of the built environment - such as uneven sidewalks, inaccessible housing, and transportation barriers - emerged as salient constraints on mobility, clinic attendance, and social participation. These constraints were noted across neighborhood contexts, indicating that structural barriers may affect aging well irrespective of neighborhood income. Differences across neighborhoods were more apparent in social and economic domains. Participants in less affluent neighborhoods described social cohesion and mutual support as important resources that buffered economic and healthcare constraints, whereas participants in more affluent neighborhoods more often emphasized financial resources as supportive. Health care access was described as both enabling and challenging across settings, reflecting structural complexity rather than a uniform advantage tied to neighborhood affluence. Together, these findings suggest that neighborhood conditions and social support networks may influence how successfully patients implement HF management strategies in daily life. Integrating social risk screening, transportation assistance, and community partnerships into HF programs may help align physiologic management with patients’ lived realities.

This study should be interpreted considering several limitations. As a qualitative, hypothesis-generating study, findings are not intended to be generalizable. Although the study focused on older adults with HF, one participant in her 40’s years old; this individual was retained because she met all other eligibility criteria, resided within a neighborhood of interest, and helped maintain balance between affluent and less affluent neighborhoods. We acknowledge that ZIP code-level socioeconomic measures may obscure within-neighborhood heterogeneity and do not capture individual-level socioeconomic experiences. However, our analytic approach was intentionally focused on contextual and structural exposures rather than individual socioeconomic status alone. ZIP code-level indicators capture shared environmental, institutional, and resource-related conditions that are central to place-based theories of health and aging. In addition, the exclusion of individuals with advanced HF, nursing home residents, hospice patients, and those with dementia limits insight into aging well among individuals with the greatest functional and cognitive impairment. As a result, our findings may overrepresent the perspectives of community-dwelling adults who are sufficiently healthy to participate in interviews, introducing potential selection bias and limiting understanding of aging well across the full spectrum of HF severity and late-life experiences. Finally, participants’ perceptions may have been influenced by response shift, whereby individuals adapt to chronic illness by recalibrating their expectations and definitions of well-being. Indeed, many participants described compensatory strategies and adaptation to functional losses, making it difficult to distinguish between successful adaptation and changing internal standards. Future research should test this proposed framework in larger and more diverse samples, incorporate longitudinal designs, and examine how functional capacity, pain, well-being, adaptation, and place-based factors interact over time to shape aging trajectories among individuals living with chronic disease.

## CONCLUSION

Among older adults living with HF, aging well is defined not solely by disease control or preserved cardiac function but by maintaining physical and cognitive capacity, sustaining social engagement, minimizing pain, and fostering overall well-being. Despite living with a chronic and often debilitating cardiovascular condition, many individuals perceive themselves as aging well. These findings expand traditional successful aging frameworks and underscore the need for HF care models that incorporate pain management, social context, and patient-defined priorities alongside conventional cardiovascular metrics. Aligning HF care with what patients value may improve patient-centered outcomes and support more comprehensive, context-sensitive cardiovascular practice.

## Data Availability

The data underlying this study consist of qualitative interview transcripts that contain potentially identifiable participant information and therefore are not publicly available. De-identified data may be made available from the corresponding author upon reasonable request and subject to institutional review board approval and applicable data use agreements.

## Acknowledgements

The authors extend their heartfelt gratitude to the participants, Weill Cornell Medicine physicians and staff who assisted with recruiting, and the Master’s Program in Clinical Epidemiology and Health Services Research who also assisted in guiding this study.

## Authors’ contribution

**Conceptualization:** Ro-Jay Reid, Parag Goyal, Atami De Main, W. Marcus Lambert, Monika M. Safford

**Data curation:** Ro-Jay Reid, Deborah Ofosu, Atami De Main, Iris Navarro-Millán, Monika M. Safford

**Formal analysis:** Ro-Jay Reid, Deborah Ofosu, Atami De Main, Iris Navarro-Millán, Madeline R. Sterling, Monika M. Safford

**Funding acquisition:** Ro-Jay Reid, W. Marcus Lambert, Monika M. Safford

**Investigation:** Ro-Jay Reid, Deborah Ofosu,

**Methodology:** Ro-Jay Reid, Deborah Ofosu, Parag Goyal, Mangala Rajan, Orysya Soroka

**Project administration:** Ro-Jay Reid, Deborah Ofosu,

**Resources**: Ro-Jay Reid, Parag Goyal, Monika M. Safford

**Supervision:** Parag Goya, Atami De Main, W. Marcus Lambert, Iris Navarro-Millán, Madeline R. Sterling, Monika M. Safford

**Writing - review & editing**: Ro-Jay Reid, Deborah Ofosu, Parag Goyal, Atami De Main, W. Marcus Lambert, Iris Navarro-Millán, Madeline R. Sterling, Mangala Rajan, Orysya Soroka, Monika M. Safford

## ABRREVIATIONS LIST

HF: Heart failure
NYHA: New York Heart Association
WHO: World Health Organization

